# Retrospective Wastewater Based Tracking of Measles Outbreak in Canton of Vaud, Switzerland: January – March 2024

**DOI:** 10.1101/2025.03.11.25322836

**Authors:** Charles Gan, Melissa Pitton, Jolinda de Korne-Elenbaas, Ludovico Cobuccio, Alessandro Cassini, Christoph Ort, Timothy R. Julian

## Abstract

Measles outbreaks remain a significant public health challenge despite high vaccination coverage in many regions. Wastewater-based surveillance (WBS) offers a non-invasive and community-level approach to monitoring the circulation of pathogens, including the measles virus. Here, we retrospectively applied a duplex digital PCR assay to distinguish between wild-type and vaccine strains of measles virus in wastewater samples available from an existing national WBS program. Samples originated from the wastewater treatment plant serving the Lausanne city catchment area, where an outbreak occurred before spreading to the broader Canton Vaud region. Despite high vaccination rates, viral loads of measles wild-type were detected during the first transmission event involving 21 cases identified within a week. However, viral loads were no longer detectable after the initial 21 cases, despite an additional 30 cases reported in the following three weeks, possibly due to lower incidence rate or location outside the catchment. Measles vaccine strain was not detected during the outbreak. Our results demonstrate the complementarity of WBS to clinical surveillance and suggest its potential as an early warning system for measles and other vaccine-preventable diseases. Further improvements in assay sensitivity and integration with epidemiological data could enhance the utility of WBS for outbreak detection and control.

**Synopsis:** Demonstrating wastewater surveillance as a complementary tool for measles outbreak monitoring, enhancing public health responses in immunized populations.

## Introduction

Measles is a highly contagious disease caused by the measles virus that can lead to complications such as pneumonia, encephalitis, and, in some cases, death^1^. Measles virus is characterized by a basic reproduction number (R0) estimated to be around 15.7, significantly higher than that of many other infectious diseases^2^. This high R0 underscores the rapidity with which the virus can spread in susceptible populations, making effective control measures crucial. A safe and highly effective vaccine has existed since the 1960s^3^. However, measles remains a public health concern in many parts of the world due to waning vaccination coverage^4^. Further, there have been multiple instances of vaccine breakthrough^5^ where people are infected despite previous vaccination. This breakthrough allows outbreaks to occur despite high vaccine coverage (>95% for herd immunity^6^). A recent example was observed in the Canton of Vaud (Switzerland)^7^, where the two-dose vaccination rate was high (91% for 2-year-olds, 94% for 8-year-olds, and 96% for 16-year-olds in 2023^8^), but still 51 confirmed cases emerged, of which 73% were vaccinated^7^.

Wastewater-based surveillance (WBS) has surfaced as a transformative tool for public health, providing an innovative approach to monitor pathogens at the community level. By analyzing wastewater for genetic material from viruses, WBS offers a cost-effective and non-invasive means to detect and track disease spread in alignment with reported clinical cases. This methodology has gained significant attention during the COVID-19 pandemic, where it proved instrumental in monitoring SARS-CoV-2 trends in real time. Beyond acute and novel outbreaks, WBS has also demonstrated utility in tracking endemic pathogens and providing early warnings for re-emerging diseases (e.g. polio)^9,10^. Furthermore, WBS could help detect breakthrough cases of diseases like measles^11^, which, although rare, can occur even in highly vaccinated communities due to waning immunity or other factors.

Measles virus has been shown to be detectable in wastewater across multiple studies^12–14^, particularly in catchments with reported clinical cases^12,13^ or symptomatic individuals^14^. WBS of measles has also demonstrated its utility in populations with predominantly low vaccination coverage^12^ as well as those with more variable vaccination statuses^13,14^. These findings highlight the potential of WBS to monitor measles virus circulation and track disease dynamics within communities. Furthermore, detection of strain-specific mutations with digital PCR (dPCR) enables differentiation between measles virus shed from community infections and that shed after vaccination, further enhancing the precision of wastewater surveillance.

From January through March 2024, a measles outbreak initiated in Lausanne city (within Canton Vaud, Switzerland) led to 51 reported clinical cases, of which at least 31 were breakthrough cases amongst people previously vaccinated with two doses^7^. Coincident analyses of wastewater for circulating endemic respiratory pathogens^15^, allowed retrospective analysis of archived wastewater extracts for detection and quantification of measles. A duplex dPCR assay targeting and differentiating between wild type (WT) measles and measles vaccine (VA) was applied to evaluate WBS for tracking dynamics of measles virus circulation in the community. This approach provides a model for integrating wastewater data into measles control strategies.

## Material and Methods

### Wastewater Composite Samples

Corresponding to the most recent measles outbreak which originated in Lausanne city and spread within the Canton of Vaud region between January and March 2024, sixty-four 24-hr composite influent samples from the wastewater treatment plant (STEP Vidy) covering the entire Lausanne city catchment area (population: 240,000) were measured for measles WT and VA between January 2^nd^ and March 31^st^, 2024 (Table S1). All samples were collected in the context of an ongoing campaign to monitor respiratory viruses in Swiss wastewater^15^. Samples were stored at 4°C and then extracted within one week of collection (2-7days) using the Wizard Enviro Total Nucleic Acid Extraction Kit (Promega, Wisconsin, USA) protocol with OneStep PCR Inhibitor Removal Kit (Zymo Research, California, USA) as described previously^16^. Extracts were stored at -80°C and analyzed between October 8^th^, 2024, and December 5^th^, 2024. Measured extracts were three-fold diluted prior to dPCR analysis to minimize inhibition.

### Digital PCR Assay

A duplex dPCR assay was developed for measles virus, enabling simultaneous differentiation and quantification of WT and VA strains. Primers and probe sequences were adapted from Wu et al.^17^ (Table S2) and were manufactured at Integrated DNA Technologies (IDT; Iowa, USA). Fluorophore for the measles WT probe was modified to contain a Freedom Cy®5.5 dye and both measles WT and VA probes were modified to contain an Iowa Black® RQ Dark Quencher (Table S2).

PCR pre-reaction volume was 27 µl, containing 5.4 µl of sample template and 21.6 µl of mastermix. Mastermix consisted of qScript XLT 1-step RT-PCR Tough Mix (2x concentrated, Quantabio, Massachusetts, USA), 500 nanomolar (nM) forward primer, 500 nM reverse primer, 200 nM WT probe, 200 nM VA probe, and RNase free water to fill. Of the total 27 µl PCR pre-reaction, 25 µl was pipetted into the chamber of the Sapphire chip (Stilla Technologies, Villejuif, France). The chip was then placed inside of a Geode (Stilla Technologies, Villejuif, France) for partitioning and thermocycling with the following steps: partitioning for 12 min at 40°C, reverse transcription at 50°C for 1 hour, enzyme activation at 95°C for 10 minutes, then 40 cycles of denaturation at 94°C for 30 seconds and annealing/extension at 55°C for 1 minute, then finally an end cycle pressure release was added according to manufacturer’s instructions.

The measles WT/VA duplex was developed on the Naica® system 6-color digital PCR system (Stilla Technologies, Villejuif, France). The duplex assay was optimized using two types of positive controls: i) live-vaccine strain for measles (Cat No. M0210000), provided by European Directorate for the Quality of Medicines & HealthCare extracted with the QiaAmp Viral RNA MiniKit (Qiagen, Hilden, Germany) following manufacturer instructions, and ii) a synthetic DNA construct for the most recent circulating genotype of measles (i.e., B3 genotype) (gBlock®, IDT, Iowa, USA) (Table S3). Assay was first validated as a single-plex assay (Text S1) and then validated as a duplex assay (Text S2) to ensure no deviations were present when multiplexing (Figure S1). Validation consisted of testing the linearity of three 10-fold dilutions in triplicate (Figure S2).

All wastewater samples were run in duplicate and with a positive control containing both measles WT and measles VA strains (Table S3). A no-template control (NTC) was also run simultaneously and all experiments with more than two positive partitions within the NTC were rerun. Partitions were classified on Crystal Miner Software version 4.0 (Stilla Technologies) using polygon gating (Figure S2). Chambers with less than 15,000 droplets were not considered and corresponding samples were assayed again. Digital minimum information for publication of quantitative real-time PCR experiments (dMIQE) guidelines for the extracts used are mentioned elsewhere^15^.

PCR inhibition by wastewater matrix was quantified according to a method described previously^15,18^. Synthetic DNA of Measles B3 (gBlock®, IDT, Iowa, USA) was spiked in a random subset of the wastewater extracts used to generate the time series. Inhibition, expressed in percentage, was defined as one minus the ratio between the amount measured in a spiked extract divided by the amount spiked in plus the endogenous measles WT within the extract. Inhibition ranged from 0 to 100% with 0 indicating no inhibition, and 100% implying a completely inhibited sample. Samples with inhibition greater than 40% were considered inhibited and rerun at a higher dilution.

### Assay Sensitivity

We first defined a sample as positive or above the Limit of Blank (LoB) (i.e. target present with 95% confidence that it is not random noise) if there were three or more positive partitions (i.e., droplets), which corresponds to approximately 5 genome copies per reaction, as described previously by Nyaruaba et al. (2022)^19^. Limit of Detection (LoD) and Quantification (LoQ) were identified by measuring decreasing concentrations with dPCR and by model fitting^20^. LoD, representing an assessment of the sensitivity of our assay, was defined as the lowest concentration above LoB at which a sample would be detected with 95% confidence. The LoD was determined empirically using multiple replicates of dilutions at low concentrations and fitting a dose-response model adapted for qPCR as described by Klymus et al. (2020)^20^, where fit conditions were set to “best” or the model with the lowest residual error. All wastewater extracts were run in duplicate, reflecting a 2-replicate LoD for this study (Text S3). The LoQ, which is the concentration at which the quantitative estimate of a concentration is a reliable estimate, was defined here as concentration at which the coefficient of variation (CoV) was 30%, in line with recommendations from literature^21,22^. To determine the LoQ, multiple replicate dilutions at moderate-to-low concentrations were measured, with the resultant data fit to a power model, 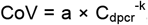, where CoV is the coefficient of variation, a is the scaling factor, C_dpcr_ is the concentration measured by dPCR, and k is the power relationship exponent. Both LoD and LoQ models were fitted with experimental data generated by measuring a minimum of 20 replicates at decreasing concentrations ranging from 110 to 6.4 gc/reaction for measles wild type (B3 gBlock®) and 40 to 8.4 gc/reaction for measles vaccine.

Both LoD and LoQ values were first expressed in gc/reaction and then multiplied by 1’200 (reactions/liter), which is a conversion factor to units of gc/L wastewater (gc/L_ww_). The conversion factor is specific to our nucleic acid extraction, dilution factor, and digital PCR detection protocols (Equation 1).

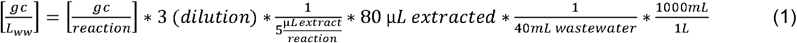

### Clinical Data

Data about clinical confirmed cases, including the number, timing and location of measles cases from people with different immunization status (fully immunized referring to 2 doses of measles-containing vaccine (MCV) or previous measles infection, partially immunized referring to 1 dose of MCV, or not immunized referring to no dose of MCV and no previous infection) was sourced from Cassini & Cobuccio et al^7^.

## Results

### Detection of Measles RNA in Wastewater Samples from Lausanne City

We applied a duplex digital PCR assay capable of differentiating measles WT and VA strains to retrospectively analyze a total of 64 wastewater samples. Of these, we detected viral loads of measles WT in nine samples (14%) that exceeded our defined LoB (Figure 1). Of these nine positive samples, two (22%) surpassed the 2-replicate LoD of 20,600 gc/L_ww_ (Figure 1). None of the positive samples exceeded the determined LoQ of 153,600 gc/L_ww_. For the quantification of measles VA, all samples were below the established LoB, determined 2-replicate limit of detection (11,100 gc/L_ww_), and determined limit of quantification (28,900 gc/L_ww_) and are omitted from further analysis. Modelled LoD and LoQ curves are provided in the supporting information (Figure S3 and Figure S4).

**Figure 1:**
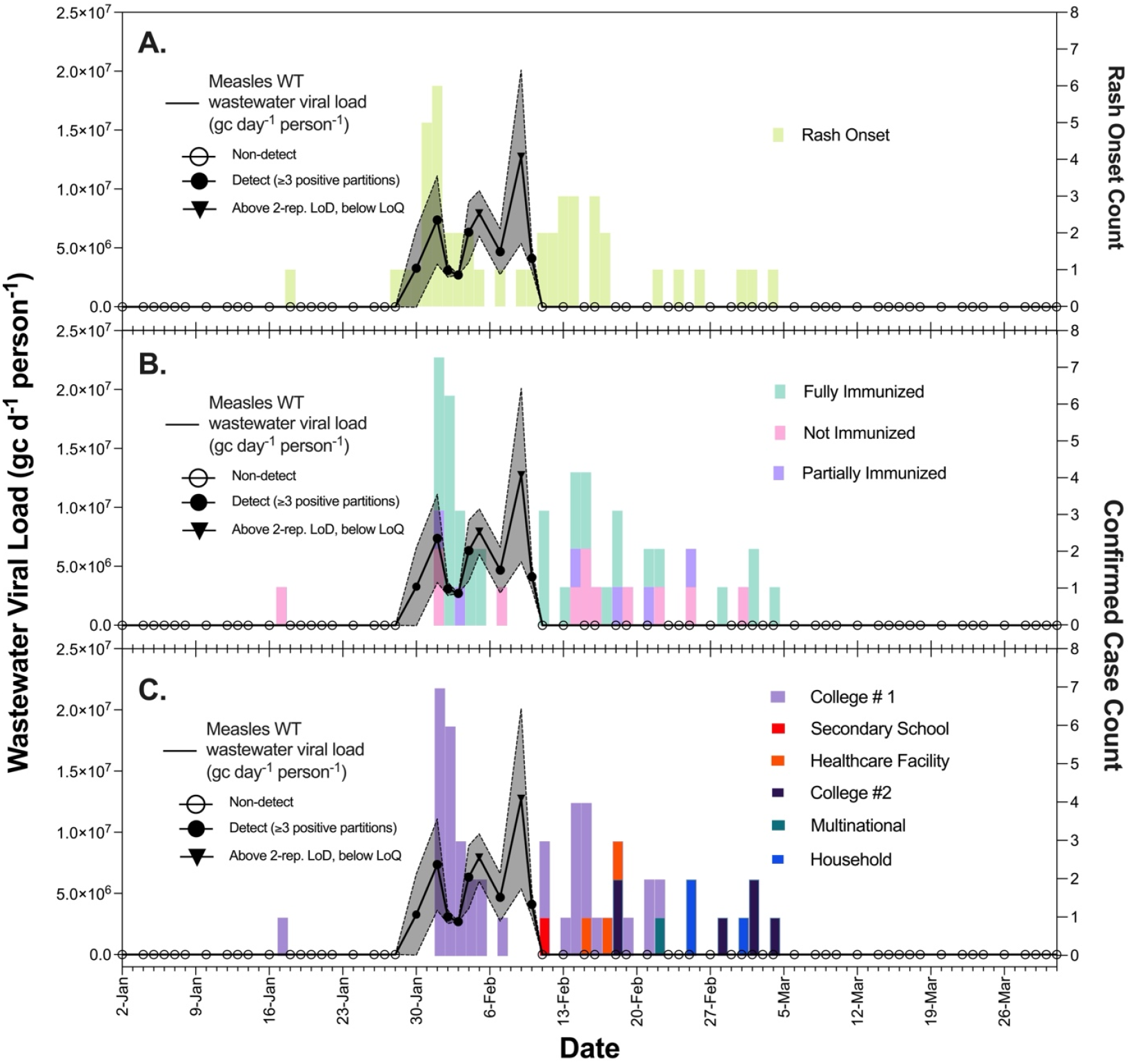
Temporal trends in measles viral load and rash onset and confirmed case counts from early January to late March 2024. **(A, B, and C)** Primary vertical axis (left) shows the viral load in wastewater expressed as gene copies per day per person (gc d^-1^ person^-1^), represented by a black line with black open circles (below LoB, i.e. non-detect), black filled circles (above LoB, i.e. 95% confidence it is not random noise; at least three positive partitions), and black inverted triangles (above 2-replicate LoD but below LoQ, i.e. detected with 95% confidence when performing two replicates, >30% expected coefficient of variation) data points. The grey shaded region indicates the standard deviation around the mean of two replicate measurements of the viral load. The horizontal axis represents the date in 2024. **(A)** The secondary vertical axis (right) shows the number of people with rash onset observed and recorded by the hospitals in the area. **(B)** The secondary vertical axis (right) shows the number of confirmed measles cases by immunization type, depicted as stacked pink (not immunized), purple (partially immunized), and green (fully immunized) bars representing immunization status. **(C)** The secondary vertical axis (right) shows the number of confirmed measles cases by location, depicted as stacked purple (college #1), red (secondary school), orange (healthcare facility), dark purple (college #2), green (multinational), and blue (household). Clinical cases, their timing, and transmission chain analysis can also be viewed at: https://www.cobuccio.me/measles_epicurve.html and https://www.cobuccio.me/transmission_chains_colorblind_proof.

To assess potential PCR inhibition, we tested a subset of 10 samples (Table S1). We observed a maximum inhibition value of 9%, which remained well below our acceptable threshold of 40%, suggesting minimal inhibition. Peaks in measles WT viral load were observed in early and mid-February, coinciding with the highest reported rash onset and case counts during the study period (Figure 1). Following mid-February, we observed a decline in viral load below our detection threshold and remained so throughout March. Notably, no rash onset events or confirmed cases were reported after March 4^th^, 2024 (Figure 1).

Overall, the viral concentrations of measles WT (averaged across duplicates) ranged from 8.25×10^3^ gc/L_ww_ to 3.3×10^4^ gc/L_ww_ (Table S1), while the corresponding measles WT viral load (averaged across duplicates) ranged from 3.09×10^6^ to 1.27×10^7^ gc day?^1^ person?^1^ (Table S1). Despite a call for vaccination issued on February 2nd, 2024^7^, we did not detect measles VA in any of the samples analyzed.

### Correspondence of Wastewater Viral Load with Rash Onset and Case Data

Rash onset, a symptom-based metric, generally preceded (likely with at least one-day lag^7^) laboratory-confirmed (98% by PCR or serology^7^) cases. Rash onset was recorded over a 46-day period from January 18^th^ to March 4^th^ with 54% (n=25) of the days having rash onset occurrences, and 22% (n=10) of days showing wastewater viral loads above the detection threshold. The detection of measles WT virus in wastewater aligned most closely with the reported rash onset occurring between January 28^th^ and February 10^th^ (14 days) consisting of 25 counts (incidence rate: 1.79 rash count per day). No wastewater viral loads were detected before or after reported rash onset counts. Additionally, no wastewater viral loads were detected in between the first case and the initial transmission cluster (January 19^th^ to January 27^th^) and from February 11^th^ to March 4^th^ in which 30 rash onset counts were recorded (incidence rate: 1.30 rash count per day).

Similarly for confirmed case counts, all case counts were recorded from January 17^th^ to March 4^th^, a total of 47 days, of which 47% (n=22) had confirmed case counts, and 21% (n=10) had wastewater loads above detection threshold. The detection of WT measles virus in wastewater aligned most closely with the 21 confirmed cases occurring between February 1^st^ and February 7^th^ (incidence rate: 3 cases per day). Like for rash onset counts, no viral load was detected immediately following the index case or in the period preceding the initial transmission cluster (January 18^th^ to January 31^st^). Furthermore, viral loads were undetectable during subsequent transmission (February 8^th^ to March 4^th^), which accounted for an additional 30 cases (incidence rate: 1.15 cases per day). Viral loads were also not detectable after no further cases were reported.

Within the initial cluster of 21 cases, 14% (n=3) were not immunized (no prior infection, no MCV dose), 10% (n=2) were partially immunized (one dose of MCV), and 76% (n=16) were fully immunized (two doses of MCV or prior infection). In contrast, the later clusters comprising 30 cases had a higher proportion of not immunized individuals (27%, n=8), with 14% (n=4) partially immunized and 59% (n=17) fully immunized. Viral loads in wastewater were most prominent during the initial cluster, which also exhibited a higher proportion of immunized individuals.

The detected measles WT viral loads in wastewater coincided with cases primarily linked to “College #1”, a confirmed location within the catchment. As viral loads became undetectable, cases associated with “College #1” declined, while the geographic distribution of cases broadened. Subsequent cases emerged from a secondary school (1 case), a healthcare facility (3 cases), “College #2” (6 cases), a multinational company (1 case), and a household (3 cases). These later cases were dispersed across western Switzerland. Due to privacy constraints, it was not possible to confirm whether these locations fell within the catchment area monitored.

## Discussion

### Detection of Measles in Wastewater and Timing of Outbreak Dynamics

Our findings demonstrate the utility of WBS for tracking transmission of measles WT virus. Detectable viral loads in wastewater were found during the first major transmission cluster of observed clinical cases, aligning with the period of highest rash onset, case confirmation, and exposure (3,700 exposed people in January 2024, then decreased afterwards from Feb 2, 2024, due to campus closure^7^). This suggests WBS could provide a complementary warning signal to case and rash data for emerging outbreaks or outbreaks in areas with no or low clinical-based surveillance.

Furthermore, clinical rash and case data are influenced by lags in reporting. Wastewater could give insight into gaps in reporting. Rash onset, a symptom-based metric, consistently preceded case confirmation by one day, a lag attributable to the time required for laboratory testing^7^. Furthermore, measles has a long incubation period of 7–14 days^1^. The decoupling of transmission events, symptom onset, and case confirmation highlights the potential of WBS as a complementary tool for bridging delays in traditional surveillance systems.

### Limitations in Detecting Extended Transmission Chains

While WBS was effective in capturing earlier cases at the start of the outbreak, WBS did not detect viral loads in the second half of the outbreak, which included 30 cases over three weeks. This may be attributed to a combination of factors, including lower exposure rates, mobility of individuals outside of the wastewater treatment plant catchment, and/or lower shedding amongst people infected later in the outbreak. From transmission chain analysis, we observed distinct clusters of cases from February 11^th^, 2024, which may have occurred outside of the surveyed catchment^7^. These results indicate that while WBS excels at identifying large-scale, early outbreak dynamics, it may be less sensitive for detecting smaller, contained transmission events which potentially span multiple catchment areas not covered by the wastewater treatment plant monitored. This underscores the need for future monitoring efforts to incorporate surrounding catchments potentially impacted by outbreaks, improving response efforts and capturing cases that stray from the initial points of transmission. Additionally, these findings highlight the importance of pairing WBS with traditional surveillance for comprehensive outbreak monitoring.

### Implications of Vaccination on Wastewater Viral Loads

The viral loads were detected during the initial cluster, which had a greater proportion of fully immunized individuals (76%), than was observed later in the outbreak (59%). Although vaccination typically reduces disease severity and viral load^5,23^, the occurrence of breakthrough cases during the initial cluster as well as the higher incidence rate may reflect higher initial attack rates which could lead to increased shedding. In contrast, later transmission clusters, with a higher proportion of not immunized individuals (27%), were not associated with detectable wastewater viral loads. We attribute this to the smaller scale of transmission and lower case and rash onset incidence rate. These findings suggest that cases amongst immunized individuals can still contribute to community-level shedding, emphasizing the need for further research to quantify shedding dynamics across vaccination statuses. While studies have characterized measles virus shedding after vaccination in individuals who do not develop the disease^24–26^, shedding patterns in vaccinated individuals with breakthrough infections remain less well understood. Understanding how factors such as the number of vaccine doses influence shedding could provide valuable insights into the detectability of measles virus in wastewater-based surveillance and its potential role in monitoring transmission dynamics.

### Outlook and Limitations

Our study underscores the potential of WBS to provide insights into measles outbreaks, complementing traditional measles surveillance or offering an alternative surveillance approach when clinical surveillance is limited or absent. Notably, the minimum threshold of cases required for reliable detection remains uncertain, as evidenced by the undetectable viral loads during reduced case incidence rate. Furthermore, spatial factors, including residence and mobility within and between wastewater treatment catchment areas, may complicate wastewater detection. More detailed demographic and mobility data may help better elucidate the relationships between clinical and wastewater-based surveillance. Finally, while the use of dPCR allowed for sensitive and specific detection of measles WT virus, we did not detect VA virus in this study despite reported vaccination campaigns. The inability of dPCR to quantify low viral loads underlines the potential of technological or methodological advancements to improve sensitivity. Future implementation of WBS for measles could enhance surveillance strategies but would benefit from improved integration with geospatial and epidemiological data, including vaccination and shedding rates. Such an approach could further improve outbreak detection and management, including potential to develop predictive frameworks.

## Data Availability

All data produced in the present study are available upon reasonable request to the authors. Data on wastewater viral loads are included in the supplemental material. Clinical case, timing, and transmission chain analysis are additionally made publicly available here: https://doi.org/10.2807/1560-7917.ES.2024.29.22.2400275. Figures representing clinical case data are made public here: https://www.cobuccio.me/measles_epicurve.html and https://www.cobuccio.me/transmission_chains_colorblind_proof.

https://www.cobuccio.me/transmission_chains_colorblind_proof

https://www.cobuccio.me/measles_epicurve.html

## Funding

This study was supported by the Swiss National Science Foundation (SNSF Sinergia Grant No. CRSII5_205933) and a grant from the Swiss Federal Office of Public Health awarded to CO and TRJ.

## Author Contributions

**Charles Gan:** Conceptualization, Methodology, Formal Analysis, Investigation, Data Curation, Writing – Original Draft, Visualization, Writing – Review and Editing. **Melissa Pitton:** Conceptualization, Formal Analysis, Writing – Original Draft, Writing – Review and Editing. **Jolinda de Korne-Elenbaas:** Conceptualization, Writing – Review and Editing. **Ludovico Cobuccio:** Data Curation, Writing – Review and Editing. **Alessandro Cassini:** Data Curation, Writing – Review and Editing. **Christoph Ort:** Conceptualization, Resources, Writing – Review and Editing, Supervision, Funding Acquisition. **Tim Julian:** Conceptualization, Methodology, Resources, Writing – Review and Editing, Supervision, Funding Acquisition.

## Conflict of Interest

None.

## Data availability

All data produced in the present study are available upon reasonable request to the authors. Data on wastewater viral loads are included in the supplemental material.

Clinical case, timing, and transmission chain analysis are additionally made publicly available here: https://doi.org/10.2807/1560-7917.ES.2024.29.22.2400275.

Figures representing clinical case data are made public here: https://www.cobuccio.me/transmission_chains_colorblind_proof and https://www.cobuccio.me/measles_epicurve.html.

## Use of LLMs

Chat-GPT 4 was used for refinement in grammar, outlining discussion, and structuring introduction.

## Notes

### Competing Interest Statement

The authors have declared no competing interest.

### Author Declarations

The study used openly available clinical data originally published here: https://doi.org/10.2807/1560-7917.ES.2024.29.22.2400275

