## Supplemental Material for "Retrospective Wastewater Based Tracking of Measles Outbreak in Canton of Vaud, Switzerland: January – March 2024"

### Text

**Text S1. Single-plex assay performance**

Prior to multiplexing, we tested each target in single-plex assays to ensure the separation between positive and negative clusters, and we tested the linearity using synthetic positive controls. Each single-plex assay showed clear distinction between positive and negative clusters, and data was fitted to linear regression, leading to R^2^ values equal or greater than 0.99 for each target (Figure S2).

**Text S2. Comparison of single-plex to duplex assays performance**

To ensure that primer and probe interactions were minimal, quantification of positive material was compared between single-plex and duplex for both duplex assays (Figure S1). To accomplish this, a mix of measles B3 strain synthetic DNA and measles vaccine was 10-fold diluted three times and then measured in single-plex and duplex for both duplexes in technical replicates. The concentrations of individual replicates were averaged, and paired t-tests were performed to determine whether significant difference existed between single-plex and duplex concentrations for each dilution. Significance was set to p = 0.05. We observed that concentrations did not significantly differ between single-plex and duplex for measles WT/VA (Figure S1) (p > 0.05 for each pairwise comparison).

**Text S3. LoD of duplex assays**

The limit of detection (LoD) is influenced by the number of replicates analyzed per sample, as the chances of detection improve with a higher number of replicates. Consequently, LoD values determined for a single replicate are extended to estimate those for varying numbers of replicates (e.g., one, two, three, four, five, and eight replicates) as shown in Figure S3. The detection probability associated with each replicate count was expressed as a single-replicate detection probability by calculating the nth-root of the non-detection probability (e.g., for 95% detection: 1 - (0.05)^(1/n)^, where n represents the number of replicates).

### Tables

**Table S1. Measles wild type RNA concentration, viral load, and inhibition in Lausanne wastewater.** C: concentration. AVG: mean. MSLS: measles. STD: standard deviation. WT: wild type. “-“: not measured.

| **Date** | **Flow Rate (m^3^)** | **C_AVG_ MSLS WT [gc/L_ww_]** | | **C_STD_ MSLS WT [gc/L_ww_]** | | **AVG of MSLS WT load (gc/d/person)** | | **STD of MSLS WT load (gc/d/person)** | **Inhibition (%)** |
| --- | --- | --- | --- | --- | --- | --- | --- | --- | --- |
| 2024-01-02 | 233900 | 0 | 0 | | 0 | | 0 | | - |
| 2024-01-04 | 118687 | 0 | 0 | | 0 | | 0 | | - |
| 2024-01-05 | 160970 | 0 | 0 | | 0 | | 0 | | - |
| 2024-01-06 | 117133 | 0 | 0 | | 0 | | 0 | | - |
| 2024-01-07 | 112260 | 0 | 0 | | 0 | | 0 | | - |
| 2024-01-08 | 101322 | 0 | 0 | | 0 | | 0 | | - |
| 2024-01-10 | 108052 | 0 | 0 | | 0 | | 0 | | - |
| 2024-01-12 | 93961 | 0 | 0 | | 0 | | 0 | | - |
| 2024-01-13 | 91343 | 0 | 0 | | 0 | | 0 | | - |
| 2024-01-14 | 93173 | 0 | 0 | | 0 | | 0 | | - |
| 2024-01-16 | 106184 | 0 | 0 | | 0 | | 0 | | - |
| 2024-01-18 | 371137 | 0 | 0 | | 0 | | 0 | | - |
| 2024-01-19 | 167938 | 0 | 0 | | 0 | | 0 | | - |
| 2024-01-20 | 129327 | 0 | 0 | | 0 | | 0 | | - |
| 2024-01-21 | 117022 | 0 | 0 | | 0 | | 0 | | - |
| 2024-01-22 | 208803 | 0 | 0 | | 0 | | 0 | | - |
| 2024-01-24 | 121704 | 0 | 0 | | 0 | | 0 | | 3 |
| 2024-01-26 | 105482 | 0 | 0 | | 0 | | 0 | | - |
| 2024-01-27 | 98898 | 0 | 0 | | 0 | | 0 | | -4 |
| 2024-01-28 | 97311 | 0 | 0 | | 0 | | 0 | | - |
| 2024-01-30 | 93713 | 8.40E+03 | 8.49E+03 | | 3.28E+06 | | 3.31E+06 | | - |
| 2024-02-01 | 92045 | 1.92E+04 | 9.76E+03 | | 7.36E+06 | | 3.74E+06 | | 3 |
| 2024-02-02 | 89750 | 8.25E+03 | 1.48E+03 | | 3.09E+06 | | 5.55E+05 | | - |
| 2024-02-03 | 86453 | 7.50E+03 | 4.24E+02 | | 2.70E+06 | | 1.53E+05 | | -4 |
| 2024-02-04 | 85837 | 1.77E+04 | 7.21E+03 | | 6.33E+06 | | 2.58E+06 | | - |
| 2024-02-05 | 87525 | 2.18E+04 | 5.30E+03 | | 7.93E+06 | | 1.93E+06 | | 2 |
| 2024-02-07 | 110160 | 1.02E+04 | 4.24E+03 | | 4.68E+06 | | 1.95E+06 | | - |
| 2024-02-09 | 92602 | 3.30E+04 | 1.91E+04 | | 1.27E+07 | | 7.37E+06 | | 8 |
| 2024-02-10 | 96657 | 1.02E+04 | 2.55E+03 | | 4.11E+06 | | 1.03E+06 | | - |
| 2024-02-11 | 96108 | 0 | 0 | | 0 | | 0 | | -4 |
| 2024-02-13 | 85468 | 0 | 0 | | 0 | | 0 | | - |
| 2024-02-15 | 82418 | 0 | 0 | | 0 | | 0 | | -2 |
| 2024-02-16 | 81992 | 0 | 0 | | 0 | | 0 | | - |
| 2024-02-18 | 78897 | 0 | 0 | | 0 | | 0 | | -1 |
| 2024-02-19 | 95700 | 0 | 0 | | 0 | | 0 | | - |
| 2024-02-21 | 82531 | 0 | 0 | | 0 | | 0 | | 9 |
| 2024-02-23 | 231231 | 0 | 0 | | 0 | | 0 | | - |
| 2024-02-24 | 111999 | 0 | 0 | | 0 | | 0 | | - |
| 2024-02-25 | 92632 | 0 | 0 | | 0 | | 0 | | - |
| 2024-02-27 | 106912 | 0 | 0 | | 0 | | 0 | | - |
| 2024-02-29 | 95245 | 0 | 0 | | 0 | | 0 | | - |
| 2024-03-01 | 92043 | 0 | 0 | | 0 | | 0 | | - |
| 2024-03-02 | 89262 | 0 | 0 | | 0 | | 0 | | - |
| 2024-03-03 | 85238 | 0 | 0 | | 0 | | 0 | | - |
| 2024-03-04 | 84652 | 0 | 0 | | 0 | | 0 | | - |
| 2024-03-06 | 85544 | 0 | 0 | | 0 | | 0 | | - |
| 2024-03-08 | 84507 | 0 | 0 | | 0 | | 0 | | - |
| 2024-03-09 | 81314 | 0 | 0 | | 0 | | 0 | | - |
| 2024-03-10 | 86489 | 0 | 0 | | 0 | | 0 | | - |
| 2024-03-12 | 85144 | 0 | 0 | | 0 | | 0 | | - |
| 2024-03-14 | 89249 | 0 | 0 | | 0 | | 0 | | - |
| 2024-03-15 | 123496 | 0 | 0 | | 0 | | 0 | | - |
| 2024-03-16 | 87515 | 0 | 0 | | 0 | | 0 | | - |
| 2024-03-17 | 119503 | 0 | 0 | | 0 | | 0 | | - |
| 2024-03-18 | 146879 | 0 | 0 | | 0 | | 0 | | - |
| 2024-03-20 | 92613 | 0 | 0 | | 0 | | 0 | | - |
| 2024-03-22 | 93594 | 0 | 0 | | 0 | | 0 | | - |
| 2024-03-23 | 91342 | 0 | 0 | | 0 | | 0 | | - |
| 2024-03-24 | 87584 | 0 | 0 | | 0 | | 0 | | - |
| 2024-03-26 | 90254 | 0 | 0 | | 0 | | 0 | | - |
| 2024-03-28 | 112914 | 0 | 0 | | 0 | | 0 | | - |
| 2024-03-29 | 87805 | 0 | 0 | | 0 | | 0 | | - |
| 2024-03-30 | 81727 | 0 | 0 | | 0 | | 0 | | - |
| 2024-03-31 | 126624 | 0 | 0 | | 0 | | 0 | | - |

**Table S2. Measles Wild Type (WT) and Vaccine (VA) primers and probes used in this study.** WT: wild type. VA: vaccine. /5Cy55/: Freedom Cy®5.5 dye modification from IDT (Iowa, USA). /5Cy5/: Freedom Cy®5 dye from IDT (Iowa, USA). /3IAbRQSp/ - Iowa Black® RQ Dark Quencher from IDT (Iowa, USA).

| **Oligo Type** | **Sequence (5’ to 3’)** |
| --- | --- |
| Forward Primer | AATGAAAAACTGGTGTTCTACAA |
| Reverse Primer | GGTGATGCTCATATAAACAACAC |
| Probe – WT | /5Cy55/TATCCAGCGGTATCAGATTAACTGCATTGCA**/3IAbRQSp/** |
| Probe – VA | /5Cy5/TATCGAGCG**/TAO/**GTATCAGATTAACCGCATTGCA**/3IAbRQSp/** |

**Table S3. Characteristics of measles WT and VA positive controls used.**

| **Positive Material Type** | **Sequence/Genbank #** | **Details** |
| --- | --- | --- |
| Measles B3 strain gBlock® | GTCCCTGCCCCTAGGTGTTGGCAGATCCACAGCAAAACCCGAAGAACTCCTCAAGGAGGCCACTGAGCTTGACATAGTTGTTAGACGTACAGCAGGGCTCAATGAAAAACTGGTGTTCTACAACAACACTCCACTAACTCTCCTCACACCTTGGAGAAAGGTCCTGACAACAGGGAGTGTCTTCAACGCAAATCAAGTGTGCAATGCGGTTAATCTGATACCGCTGGATACCCCGCAGAGGTTCCGTGTTGTTTATATGAGCATCACCCGTCTTTCAGATAACGGGTATTACACTGTTCCTAGAAGAATGCTGGAATTCAGATCAGTCAATGCAGTGGCCTTCAACCTGCTGGTGACCCTTAGGATTG | Nucleotide 3656-4025 |
| Measles Schwarz vaccine strain | Genbank # GCA_031113345.1 | Whole Genome |

### Figures

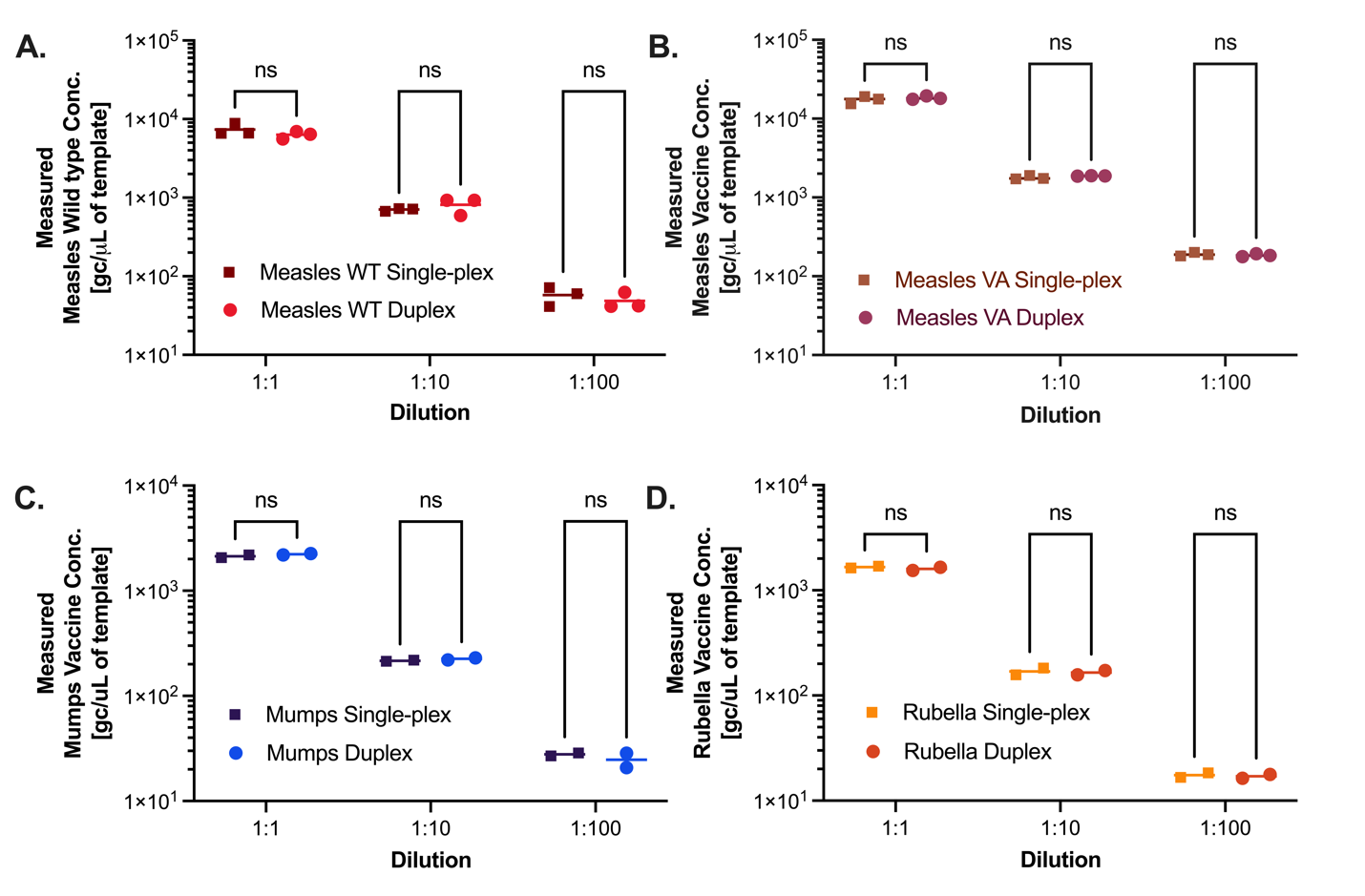

**Figure S1. Quantification comparison between single-plex assays and duplexes using positive controls.** Horizontal axes show three 10-fold dilutions of the positive material. Y-axes indicate the measured concentrations using dPCR and expressed in genome copies (gc) per microliter (µl) of template. Within each dilution category, individual squares and circles represent replicate measurements of the same sample. Viral targets are specified with distinct colors and sorted by panel: measles WT in panel A and measles vaccine in panel B. Statistical significance was inferred using multiple paired t-tests. ns: not significant. All p-values were greater than 0.24 with significance threshold established as p < 0.05.

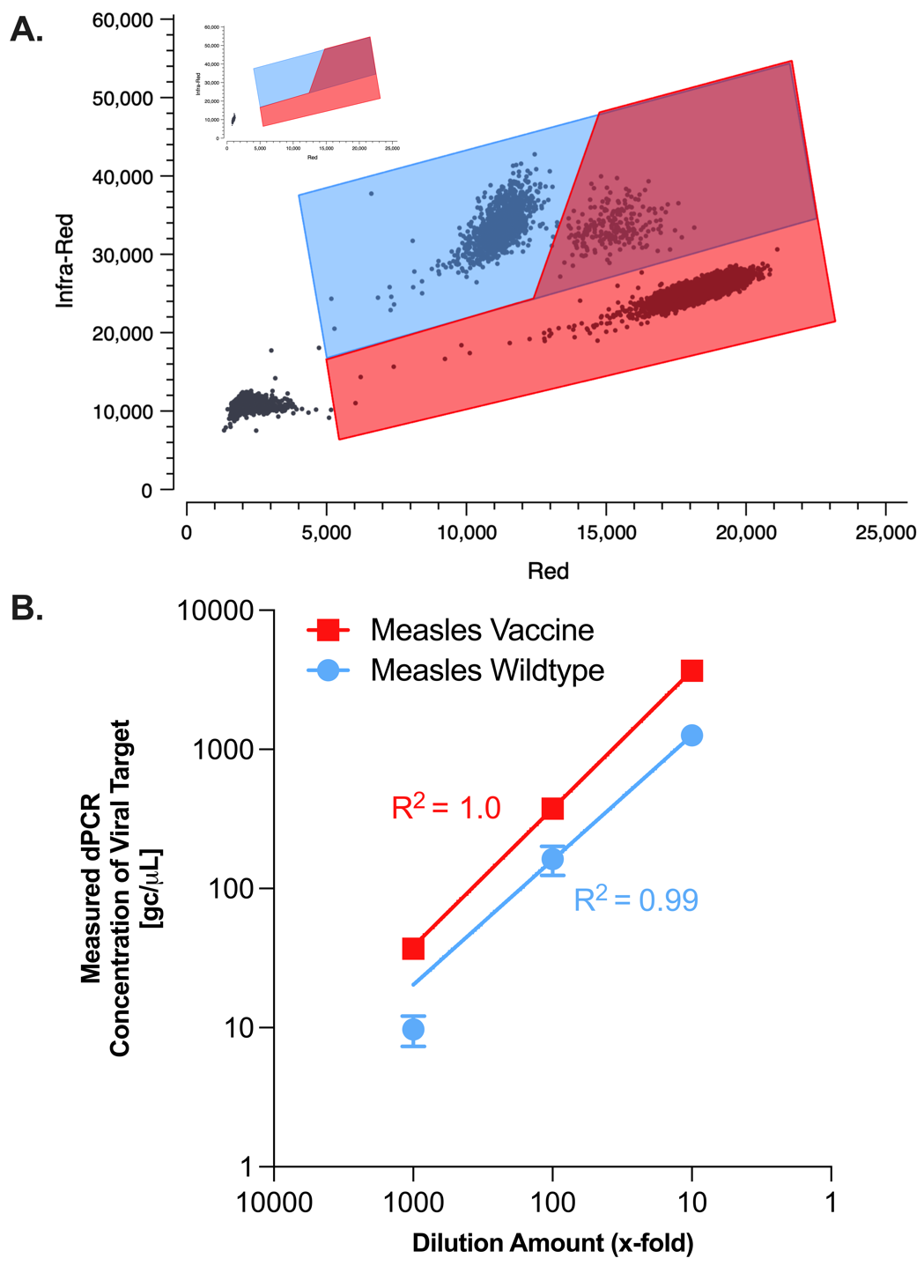

**Figure S2. Validation of single-plex dPCR assays.** (A) Two dimensional representations of partition classification. Horizontal axis shows fluorescent intensity in the red channel. Vertical axis displays the fluorescent intensity in the infrared channel. Each dot represents a single partition. Positive partitions are classified by polygon gating where the blue polygon contains partitions with measles WT and the red polygon contains partitions with measles VA. (B) Linearity testing for duplex assay. Red line represents the concentration of measles VA and blue line represents the concentration of measles WT. Horizontal axis indicates the fold dilution factor (10-fold dilutions). Vertical axis illustrates the measured concentration using dPCR and it is expressed in genome copies (gc) per microliter (µl) of template. Each dilution was measured in technical triplicates. Data was fitted to linear regression and R^2^ values are displayed for each target (R^2^ = 1.0 for measles VA and R^2^ = 0.99 for measles WT).

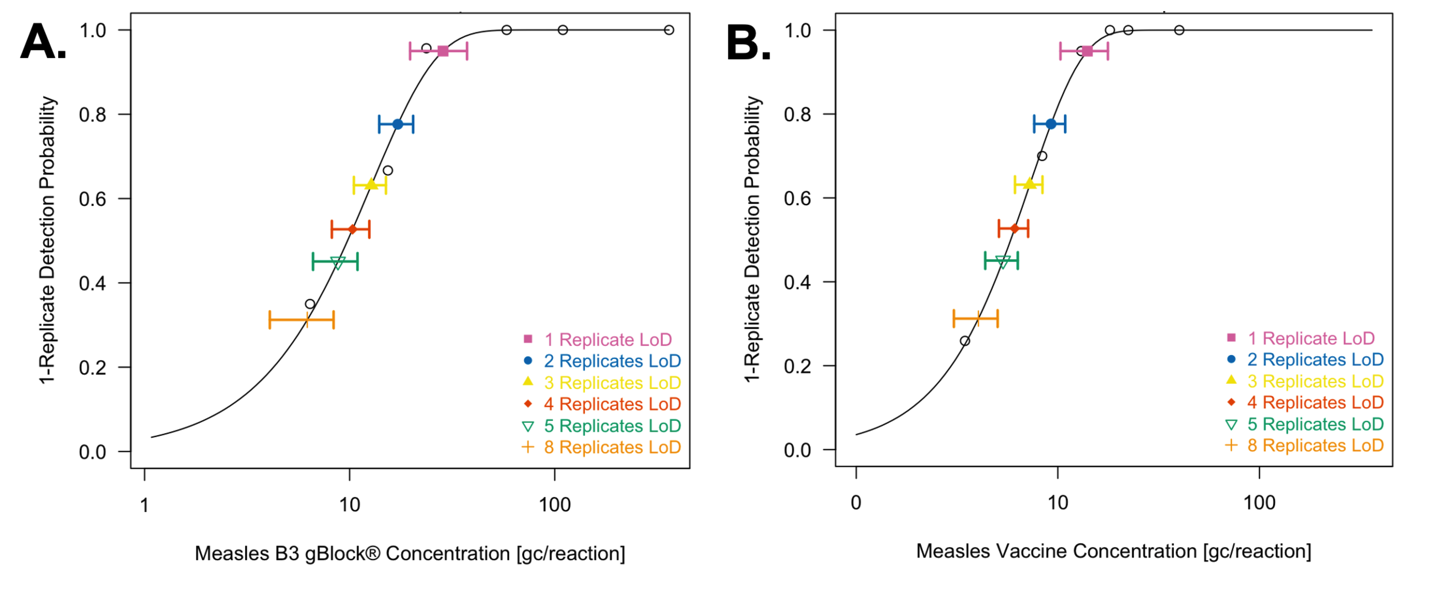

**Figure S3. Modelled LoD of all targets measured using measles WT and VA duplex assay.** Weibull Type-2, two-parameter detection probability curves show the likelihood of detecting each viral target at increasing concentrations, expressed as genome copies (gc) per reaction. Panels A and B represent different viral targets: (A) measles B3 gBlock® concentration and (B) measles VA concentration. Each colored line and symbol represent the limit of detection (LoD) for different numbers of replicates (1 to 8). The probability of detection approaches 1.0 as the target concentration increases, with LoD improving (shifted left) as replicate numbers increase. Error bars indicate the asymptotical Wald-type confidence intervals for each LoD threshold.

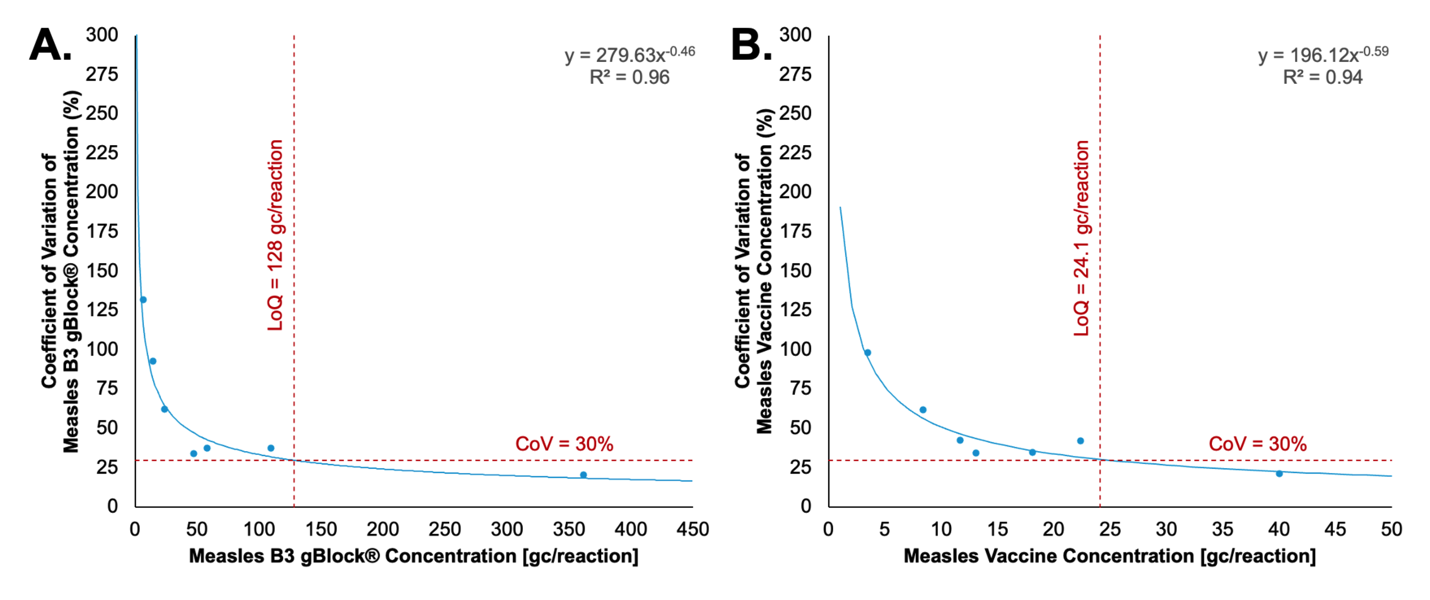

**Figure S4. Modelled LoQ of all targets measured using measles WT and VA duplex assay.** (A and B) Viral target concentration is represented on the horizontal axis in gene copies per reaction. Coefficient of variation at each concentration is represented on the vertical axis. A power law model is fitted through experimental data. The panels A and B represent the different viral targets. (A) Measles B3 gBlock® concentration: LoQ at 128 gc/reaction, with a model fit equation y = 279.63x^−0.46^ and R^2^ = 0.96 (B) Measles vaccine concentration: LoQ at 24.1 gc/reaction, with a model fit equation y = 196.12x^−0.59^ and R^2^=0.94 CoV decreases as target concentration increases. The dashed horizontal red line indicates the 30% CoV threshold for reliable quantification, and the vertical dashed red line represents the interpolated LoQ for each target.
